# Bivalent booster effectiveness against severe COVID-19 outcomes in Finland, September 2022 – March 2023

**DOI:** 10.1101/2023.03.02.23286561

**Authors:** Eero Poukka, Hanna Nohynek, Sirkka Goebeler, Tuija Leino, Ulrike Baum

## Abstract

Bivalent COVID-19 vaccines were introduced in 2022 but knowledge of their effectiveness against severe COVID-19 outcomes is currently limited. In Finnish register-based cohort analyses, we compared the risk of severe COVID-19 outcomes among those who received bivalent vaccination (exposed) between September 2022 and March 2023 to those who did not (unexposed). Among elderly aged 65–110 years, bivalent vaccination reduced the risk of hospitalisation and death due to COVID-19 in September–December 2022; the hazard ratios comparing exposed and unexposed ranged from 0.37 to 0.45 during the first 31–60 days since bivalent vaccination. However, in January–March 2023 the effect disappeared possibly indicating immune evasion of new SARS-CoV-2 variants, waning of vaccine effectiveness and increased presence of hybrid immunity. Among the chronically ill aged 18–64 years bivalent vaccination did not reduce the risk of severe COVID-19 outcomes. These results are important for developing COVID-19 vaccines and programmes worldwide.

## Main text

Due to the emergence of new SARS-CoV-2 variants with immune evasive capabilities^1^, new, bivalent COVID-19 vaccines, containing mRNA that encodes the spike proteins of the original virus strain and the Omicron variant, were developed in 2022. The European Medicines Agency authorized the first (BA.1 and BA.4-5) bivalent vaccines in September 2022^2,3^, which were promptly recommended in Finland as a booster for all people aged 65 years or more and those aged 18–64 years with underlying medical conditions predisposing to severe COVID-19.

In previous studies, bivalent vaccines have increased the protection against severe outcomes^4–11^. However, the duration of this protection is currently unclear; it is expected to vary by time since vaccination and across different Omicron lineages. In studies conducted in United Kingdom and Canada, the effectiveness of bivalent vaccines was slightly reduced against sublineage BQ.1^9,12^ and possibly even further reduced against the sublineages CH.1.1 and XBB.1.5^12^. Thus, studies estimating the effectiveness of bivalent vaccines over time are needed as policy makers are considering the recommendation of a second bivalent booster. Presently, policies vary between countries: some countries decided to recommend a second bivalent booster^13^, while others do not yet recommend further boosters for spring 2023.

The aim of this study was to estimate the effectiveness of BA.1 and BA.4-5 bivalent COVID-19 vaccines against severe COVID-19 outcomes up to five months after vaccination in Finland based on national register data. Since bivalent boosters were primarily offered to individuals who had received at least two monovalent doses, our study was restricted to these vaccinees. The study period was divided into two subperiods, i.e., September–December 2022 and January–March 2023, to account for the change in circulating Omicron lineages (Supplementary Figure S1). To control for confounding, all analyses were adjusted for a set of potential confounders; a negative control outcome was used to assess the presence of residual confounding.

## Results

The study cohorts included 1,197,501 elderly aged 65–110 years and 444,327 chronically-ill individuals aged 18–64 years (Supplementary Table S3). Only a small proportion of each cohort had been laboratory-confirmed SARS-CoV-2-positive prior to the study (Supplementary Table S4). The 2022–2023 influenza vaccination coverage reached 58% in the elderly cohort and 37% in the chronically ill cohort.

During the study 645,861 (52%) elderly and 71,497 (15%) chronically ill were vaccinated with a bivalent booster; approximately a third of them received Comirnaty BA.1 while the other two thirds received Comirnaty BA.4-5. Spikevax bivalent vaccines were used only in small quantities. The median time since bivalent vaccination by the end of follow-up was 133 days (interquartile range 116–143 days) and 128 days (interquartile range 106–143 days) among the elderly and chronically ill, respectively.

Among the elderly, we observed altogether 2,013 hospitalisations due to COVID-19, 1,167 deaths due to COVID-19 and 984 deaths in which COVID-19 was a contributing factor. In September–December 2022, during the first 14–30 and 31–60 days since vaccination, a bivalent booster lowered the risk of hospitalisation due to COVID-19 (hazard ratio [HR] 0.41, 95% confidence interval [CI] 0.31–0.55; HR 0.37, 95% CI 0.27–0.50), death due to COVID-19 (0.34, 0.23–0.66; 0.47, 0.33–0.66) and death in which COVID-19 was a contributing factor (0.40, 0.27–0.60; 0.45, 0.31–0.64) (Fig. 1, Supplementary Table S5). Thereafter, in January–March 2023, the HRs were higher and ranged during days 31–150 between 0.71 (95% CI 0.46– 1.12) and 1.23 (0.79–1.91) for hospitalisation due to COVID-19, 0.75 (0.42–1.33) and 0.90 (0.44–1.87) for death due to COVID-19, and 0.40 (0.24–0.67) and 1.30 (0.63–2.66) for death in which COVID-19 was a contributing factor. Overall, in January–March 2023, the risk of severe COVID-19 outcomes was similar between those who did and those who did not receive a bivalent booster at any time since vaccination. When stratified by age, the CIs of the HRs were wide and broadly overlapping; in September-December 2022 the HRs for 65-79-year-olds and 80-110-year-olds were comparable (Fig. 2, Supplementary Tables S6– 7). Both BA.1 and BA.4–5 bivalent vaccines reduced the risk of severe COVID-19 outcomes; the HRs were similar (Supplementary Table S8).

**Figure 1.**
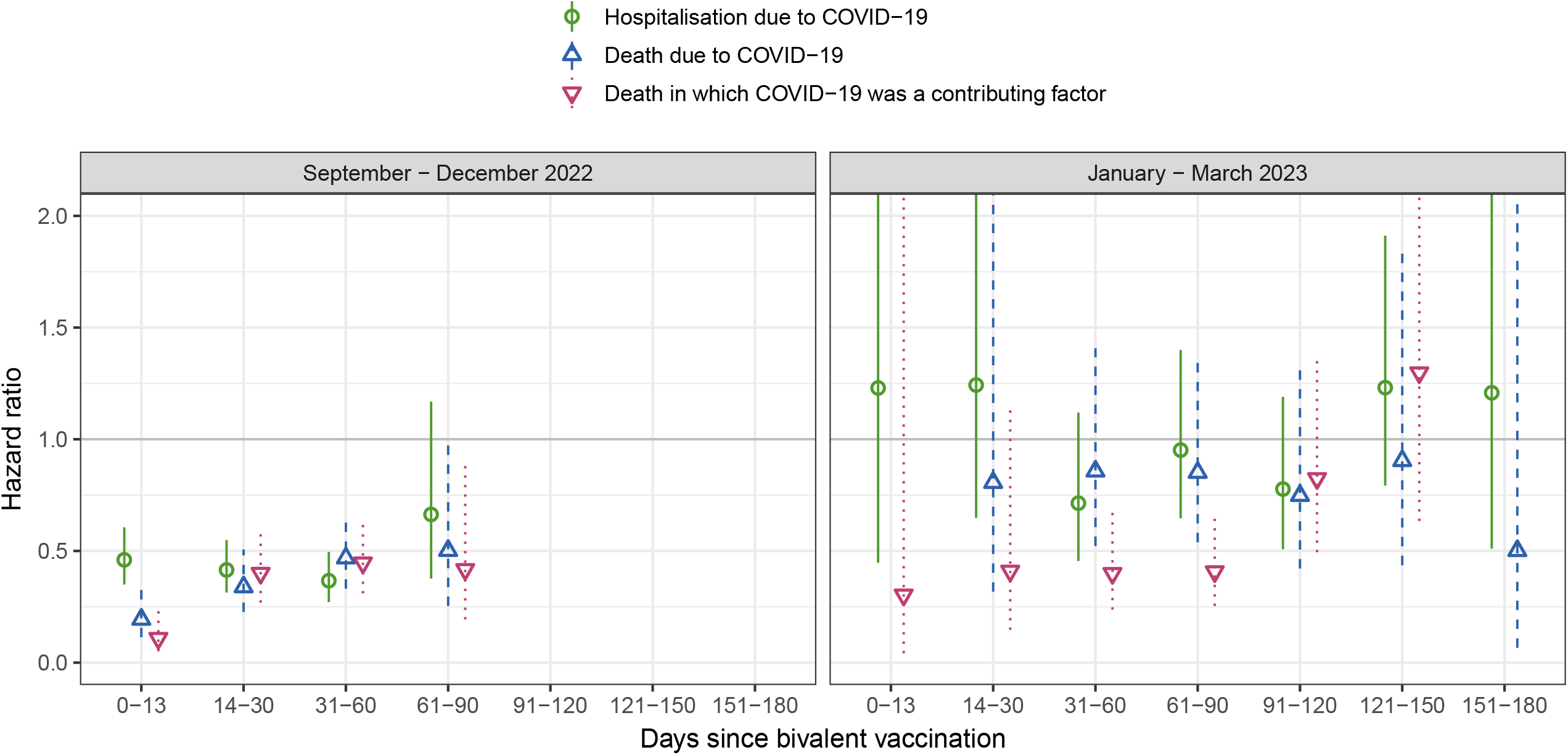
Hazard ratios among the elderly. Covariate-adjusted hazard ratios (with 95% confidence intervals) comparing the hazards of severe COVID-19 outcomes in 65-to-110-year-olds who received a bivalent COVID-19 vaccine with the corresponding hazards in those who did not receive a bivalent COVID-19 vaccine, Finland.

**Figure 2.**
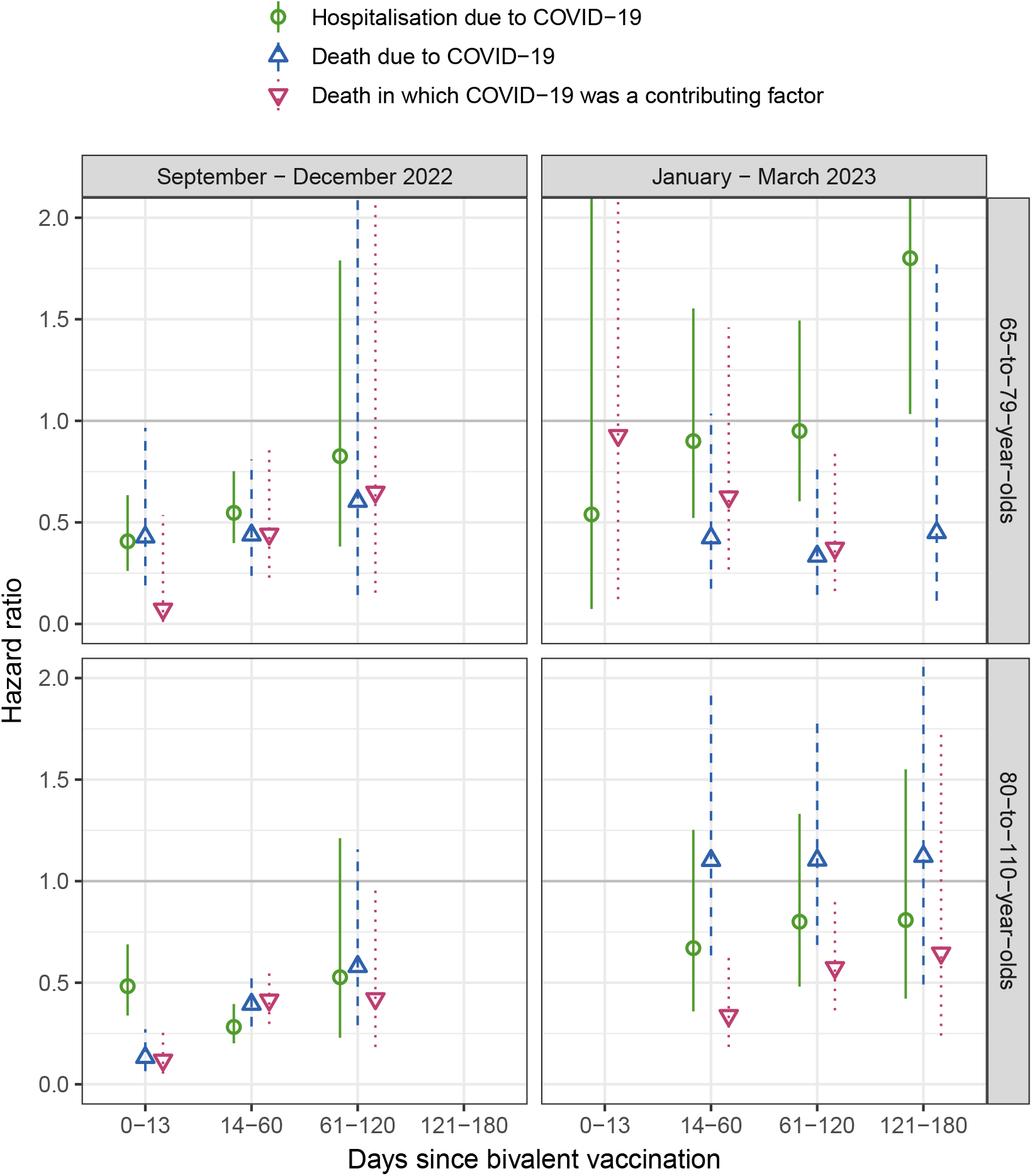
Age-stratified hazard ratios among the elderly. Covariate-adjusted hazard ratios (with 95% confidence intervals) comparing the hazards of severe COVID-19 outcomes in 65-to-110-year-olds who received a bivalent COVID-19 vaccine with the corresponding hazards in those who did not receive a bivalent COVID-19 vaccine stratified by age group, Finland.

Among the chronically ill we observed altogether 278 hospitalisations due to COVID-19, 18 deaths due to COVID-19 and 20 deaths in which COVID-19 was a contributing factor. In September–December 2022, the HR of hospitalisation due to COVID-19 was 1.01 (95% CI 0.43–2.40) for days 14–30 since bivalent vaccination and 1.81 (0.87–3.76) for the subsequent 30 days. In January–March 2023, that HR ranged between 0.74 (95% CI 0.16–3.30) and 3.40 (1.34–8.66) during days 31–150. The HR for the other two outcomes could not reliably be estimated (Supplementary Table S9).

In our negative control outcome analysis, we observed 16,577 emergency room visits due to injury among the elderly and 3,765 such visits among the chronically ill. We found no major difference in the risk of injury among elderly who received a bivalent booster and those who did not (Supplementary Table S5). However, among the chronically ill who received a bivalent booster the risk of injury appeared slightly elevated (Supplementary Table S9).

## Discussion

In our Finnish study, a bivalent booster reduced the risk of severe COVID-19 outcomes among the elderly in September–December 2022. Furthermore, bivalent boosters were equally effective among individuals aged 65–79 years and those aged 80 years or more. However, a bivalent booster appeared to neither reduce the risk of severe COVID-19 outcomes among the elderly in January–March 2023 nor benefit the chronically-ill 18–64-year-olds. For the interpretation of our findings, it is crucial to note that we estimated only the incremental effect of bivalent boosters as the comparison was not done between vaccinated and unvaccinated individuals.

We suggest three hypotheses for the reduced protective effect of bivalent boosters among elderly in January–March 2023. Firstly, at the beginning of the study BA.5 was the predominant Omicron lineage in Finland (Supplementary Figure S1), but in January–March 2023 SARS-CoV-2 infections were mostly caused by the sublineages BQ.1, CH.1.1 and XBB.1.5, which have better immune evasive capabilities than BA.5^14–17^ and have been associated with lower vaccine effectiveness estimates^9,12^. In addition, immune imprinting might have contributed to the lower protection levels observed in our study^18,19^. Secondly, the waning of effectiveness over time since vaccination, as observed in previous studies^8–10,12^, might have negatively affected the protection levels in January–March 2023. Because the potential effect of waning coincided with the emergence of new sublineages, our study does not facilitate a clean estimation of the effect of time since vaccination. To distinguish these two issues, other studies with access to viral sequencing data are required. Thirdly, the prevalence of hybrid immunity in the study population increased during the study period due to plenty of SARS-CoV-2 infections, which to a large but unknown proportion remained unregistered and, thus, could not be numerically considered in the study. As hybrid immunity provides high protection against severe COVID-19 outcomes^20^, its increased prevalence might have attenuated the additional benefit of a bivalent booster in our study by some margin.

If other studies also observe a notable reduction of bivalent vaccine effectiveness against severe COVID-19 outcomes caused by BQ.1, CH.1.1 and XBB.1.5, it signals the importance of updating the vaccine composition. This has also major programmatic implications: It might lead to withdrawal of the current BA.1 and BA.4-5 bivalent vaccines in the near future. Furthermore, it would suggest that COVID-19 vaccination campaigns must be constantly reviewed in the light of the ever-changing epidemic situation, economic analyses and vaccine supply.

Among the chronically ill we did not observe bivalent vaccination to reduce the risk of severe COVID-19 outcomes, although previous studies have found a benefit among working-age adults^6^. This may be due to several reasons. Firstly, only a small proportion of the cohort received a bivalent booster, and the negative control outcome analysis indicated the presence of residual confounding. Secondly, individuals who did not receive the booster might have had higher likelihood of unregistered SARS-CoV-2 infection and thus hybrid immunity prior to the study, which could have led to underestimation of the effectiveness. Thirdly, the number of cases among the chronically ill was small, which together with the low bivalent vaccine uptake led to unprecise estimates for that group. Fourthly, a good baseline protection due to monovalent vaccinations and hybrid immunity among the chronically ill might have limited the additional benefit of a bivalent booster.

As another limitation, we observed a decreased risk of severe COVID-19 outcomes during the first 0–13 days since bivalent vaccination. This was probably caused by selection (i.e. healthy vaccinee) bias as individuals with acute respiratory symptoms, a predeterminant of severe COVID-19 outcomes, were not advised to seek vaccination. However, it should be noted that the effect of this bias diminishes over time and is likely negligible after 13 or latest 30 days since vaccination.

Our study has also several strengths. The study was timely and representative. We used the monovalent vaccinated as the reference group, whose characteristics are probably more like the characteristics of the bivalent vaccinated compared to those of the unvaccinated. Furthermore, we did not observe major residual confounding in the negative control outcome analysis among the elderly. The recording of vaccinations and COVID-19 outcomes is mandatory, and the utilised registers have been well maintained as they have been used for routine surveillance of the COVID-19 vaccination programme and the COVID-19 pandemic in Finland.

In conclusion, bivalent boosters reduced the risk of hospitalisation and death due to COVID-19 in September–December 2022 among the elderly who had previously received at least two monovalent doses but not in January–March 2023 possibly indicating immune evasion of new SARS-CoV-2 variants, waning of vaccine effectiveness and increased presence of hybrid immunity. We did not observe an additional benefit of bivalent boosters among chronically-ill 18–64-year-olds. These findings have major vaccine development and programmatic implications, such as a need for updated COVID-19 vaccines and a refined selection of subpopulations targeted by future vaccination campaigns.

## Supporting information

Ethical statement

## Data Availability

By Finnish law, the authors are not permitted to share individual-level register data. The computing code is available upon request.

## Authors’ contributions

EP, UB, HN and TL conceptualised the study. UB conducted the statistical analysis and SG provided the death certificate data. EP and HN reviewed the literature. EP and UB drafted the manuscript. EP, UB, SG HN and TL gave comments and revised the manuscript.

## Research ethics

By Finnish law, the Finnish Institute for Health and Welfare (THL) is the national expert institution to carry out surveillance on the impact of vaccinations in Finland (Communicable Diseases Act, https://www.finlex.fi/en/laki/kaannokset/2016/en20161227.pdf). Neither specific ethical approval of this study nor informed consent from the participants was needed.

## Acknowledgements

We thank Erika Lindh and Aapo Juutinen for providing Supplementary Figure 1 as well as Heini Salo and Toni Lehtonen for the register-based identification of individuals with medical conditions predisposing to severe COVID-19. Additional thanks go to all the colleagues at the Finnish Institute for Health and Welfare (THL) who curate the register data. Lastly, we are grateful for our fruitful collaboration with Statistics Finland.

## Online Methods

We conducted population-based cohort analyses linking national register data from Finland using a unique person identifier. The study period was from 1 September 2022 to 31 March 2023, when various sublineages of Omicron were the dominant SARS-CoV-2 strains (Supplementary Figure S1). In analogy to our previous study^21^, we formed two cohorts of individuals aged 65 years or more (the elderly) and individuals aged 18–64 years with comorbidities or medical therapies predisposing to severe COVID-19 (the chronically ill, Supplementary Tables S1-2). We included only individuals that had received at least two monovalent COVID-19 vaccine doses (Comirnaty/ tozinameran/ BNT162b2, Spikevax/ elasomeran/ mRNA-1273, or Vaxzevria/ ChAdOx1-SARS-COV-2/ AZD1222). In addition, we excluded individuals that were hospitalised due to COVID-19 at the beginning of the study or had received a COVID-19 vaccination with too short dosing interval or a bivalent vaccination prior to the study (Supplementary Table S3).

The exposure was defined as vaccination with a BA.1 or BA.4-5 bivalent vaccine recorded in the Finnish Vaccination Register and was time-dependently categorized into seven groups: not vaccinated with a bivalent booster (the reference), and 0–13, 14–30, 31–60, 61–90, 91–120, 121–150, 151–180 and 181 or more days since vaccination with a bivalent booster. The bivalent vaccines included in this study were either based on Comirnaty or Spikevax as other bivalent vaccines were not available in Finland during the study period.

The severe COVID-19 outcomes were hospitalisation due to COVID-19, death due to COVID-19 and death in which COVID-19 was a contributing factor. Hospitalisations, recorded in the Care Register for Health Care, had to fulfil the following two criteria to be considered as hospitalisations due to COVID-19:

1. The primary diagnosis was COVID-19 (International Classification of Diseases, 10th revision: U07.1, U07.2), acute respiratory tract infection (J00– J22, J46) or severe complication of lower respiratory tract infections (J80–84, J85.1, J86).
2. A positive PCR-or antigen SARS-CoV-2 sample was taken from the hospitalized patient in the period extending from 14 days before to 7 days after hospital admission and registered in the National Infectious Diseases Register.

To define the two COVID-19 death outcomes, we used data collected from death certificates. In Finland, physicians record the cause of death of their patients as well as other significant conditions contributing to death in death certificates that are thereafter reviewed by medico-legal specialists at the Finnish Institute for Health and Welfare prior to forming statistics. In our study, death due to COVID-19 included all deaths in which COVID-19 was recorded as the cause of death in the death certificate. The cases in which COVID-19 was a contributing factor to death were equally retrieved from the death certificates. For the study, the data from reviewed death certificates were computerized into a database by medico-legal specialists accepting the ICD10 codes U07.1, U07.2, U09, and U10 as COVID-19 diagnosis.

In addition, we defined a fourth endpoint, which we assumed to be unaffected by the exposure. This negative control outcome was any emergency room visit due to injury (International Classification of Diseases, 10th revision: S00–T14) recorded in the Care Register for Health Care.

We considered nine covariates as confounders in our study: age group, region of residency, sex (Population Information System), hospitalisation between 1 September 2021 and 31 August 2022 (Care Register for Health Care), presence of comorbidities or medical therapies predisposing to severe COVID-19 (Care Register for Health Care, Register of Primary Health Care visits, Special Reimbursement Register for Medicine Expenses and Prescription Centre database, residency in a long-term care facility (Care Register for Social Care), seasonal influenza vaccination in 2022–2023, number of monovalent COVID-19 vaccinations (Finnish Vaccination Register) and last laboratory-confirmed SARS-CoV-2 infection prior to the study (National Infectious Diseases Register). Previous SARS-CoV-2 infections were categorised into pre-Omicron infections (before 2022) and Omicron infections (since 2022).

The individual follow-up period started earliest on 1 September 2022 and latest 91 days after the last monovalent COVID-19 vaccination or laboratory-confirmed SARS-CoV-2 infection prior to the study. Each individual was followed until death, outcome of interest, day 14 (if the outcome of interest was hospitalisation due to COVID-19) or day 60 after laboratory-confirmed SARS-CoV-2 infection, a second bivalent vaccination, a monovalent vaccination and 31 March2023, whichever occurred first.

Separately for the two calendar periods September–December 2022 and January–March 2023 and for each cohort, we compared the hazard of the three severe COVID-19 outcomes between unexposed and exposed individuals taking into account time since bivalent vaccination. The hazard ratio (HR) was estimated using Cox regression with calendar time as the underlying time scale and adjusted for the aforementioned covariates. Additionally, we stratified the elderly cohort by age differentiating between individuals aged 65–79 years and those aged 80 years or more and analysed the HR separately for BA.1 and BA.4-5 bivalent vaccines. For these analyses, we combined the second and third, fourth and fifth, and sixth and seventh time since vaccination interval to account for the expectably small number of cases in these exposure groups.

To evaluate the presence of residual confounding, we estimated the hazard ratio for the negative control outcome and expected to find no difference between the unexposed and exposed. The analysis was conducted as described above considering the negative control outcome as the outcome of interest. All analyses were performed in R 4.2.2 (R Foundation for Statistical Computing, Vienna, Austria).

