## Supplementary Information for "Bivalent booster effectiveness against severe COVID-19 outcomes in Finland, September 2022 – March 2023"

**Table S1: Highly predisposing comorbidities or medical therapies** Definition of comorbidities and medical therapies that were considered highly predisposing to severe COVID-19.

| Classification system | Codes |
|---|---|
| **Actively treated cancer** | |
| ICD10 | C00, C01, C02, C03, C04, C05, C06, C07, C08, C09, C10, C11, C12, C13, C14, C15, C16, C17, C18, C19, C20, C21, C22, C23, C24, C25, C26, C27, C28, C29, C30, C31, C32, C33, C34, C35, C36, C37, C38, C39, C40, C41, C42, C43, C45, C46, C47, C48, C49, C50, C51, C52, C53, C54, C55, C56, C57, C58, C59, C60, C61, C62, C63, C64, C65, C66, C67, C68, C69, C70, C71, C72, C73, C74, C75, C76, C77, C78, C79, C80, C81, C82, C83, C84, C85, C86, C87, C88, C89, C90, C91, C92, C93, C94, C95, C96, C97, D051, D39 |
| **Down syndrome** | |
| ICD10 | Q90 |
| **Organ or stem cell transplantation** | |
| ICD10 | T86, Z94 |
| **Severe chronic respiratory disease** | |
| ICD10 | E84, J41, J42, J43, J44, J45, J46, J47, Z902 |
| ICPC2 | R96 |
| **Severe disorder of the immune system** | |
| ICD10 | D7081, D7089, D80, D81, D82, D83, D84, E3100, E723, E76, E791, E792, E798, E799, Q7782, Q7800, Q7808, Q809, Q8900, Q8933, Q8934, Q938, Z908 |
| NCSP | JMA |
| **Severe kidney disease** | |
| ICD10 | E102, E112, E142, I12, I13, N00, N01, N02, N03, N04, N05, N07, N08, N11, N14, N18, N19 |
| **Type 2 diabetes mellitus** | |
| ATC | A10B |
| ICD10 | E11, E13, E14 |
| ICPC2 | T90 |

ATC, Anatomical Therapeutic Chemical Classification System;

ICD10, International Statistical Classification of Diseases, tenth revision;

ICPC2, International Classification of Primary Care, second edition;

NCSP, Nordic Classification of Surgical Procedures

**Table S2: Moderately predisposing comorbidities or medical therapies** Definition of comorbidities and medical therapies that were considered moderately predisposing to severe COVID-19.

| Classification system | Codes |
|---|---|
| **Autoimmune disease** | |
| ICD10 | D86, K50, K51, L40, M02, M05, M06, M07, M139, M45, M460, M461, M469, M941 |
| **Immunosuppressive medication** | |
| ATC | H02AB02, H02AB04, H02AB06, H02AB07, L01BA01, L01XC02, L04AA06, L04AA10, L04AA13, L04AA18, L04AA24, L04AA26, L04AA29, L04AA33, L04AA37, L04AB, L04AC, L04AD01, L04AD02, L04AX01, L04AX03 |
| **Neurological condition affecting breathing** | |
| ICD10 | G20, G21, G22, G23, G24, G25, G26, G70, G71, G72, G73, G80, G81, G82, G83, I60, I61, I62, I63, I64, I65, I66, I67, I68, I69 |
| **Psychotic disease** | |
| ATC | N05AH02 |
| ICD10 | F20, F21, F22, F23, F24, F25, F26, F27, F28, F29 |
| ICPC2 | P72 |
| **Severe chronic liver disease** | |
| ICD10 | K702, K703, K704, K71, K72, K73, K74 |
| **Severe heart disease** | |
| ICD10 | I110, I119, I12, I130, I131, I132, I139, I15, I20, I21, I22, I23, I24, I25, I26, I27, I28, I418, I42, I43, I50 |
| **Sleep apnea** | |
| ICD10 | G473 |
| NCSP | WX723, WX780 |
| **Type 1 diabetes mellitus or adrenal insufficiency** | |
| ATC | A10A |
| ICD10 | E10, E250, E271, E272, E274, E3100, E3101, E3108, E896 |
| ICPC2 | T89 |

ATC, Anatomical Therapeutic Chemical Classification System;

ICD10, International Statistical Classification of Diseases, tenth revision;

ICPC2, International Classification of Primary Care, second edition;

NCSP, Nordic Classification of Surgical Procedures

**Table S3: Enrollment of the study cohorts** Each target population included all individuals who were living in Finland during the study, met the age criterion at the beginning of the study, and had received at least two monovalent COVID-19 vaccine doses prior to the study.

| | No. of individuals |
|---|---|
| **65-to-110-year-olds** | |
| Target population | 1222237 |
| Excluded due to hospitalisation due to COVID-19 at the beginning of the study | 455 |
| Excluded due to too short dosing interval* prior to the study | 8656 |
| Excluded due to bivalent vaccination prior to the study | 27 |
| Excluded due to right-censoring prior to the start of individual follow-up | 15598 |
| Study cohort | 1197501 |
| **18-to-64-year-olds with underlying medical conditions** | |
| Target population | 450578 |
| Excluded due to hospitalisation due to COVID-19 at the beginning of the study | 163–166 |
| Excluded due to too short dosing interval* prior to the study | 4075 |
| Excluded due to bivalent vaccination prior to the study | 1–4 |
| Excluded due to right-censoring prior to the start of individual follow-up | 2009 |
| Study cohort | 444327 |

* less than 91 days between two doses or
less than 14 days between the first two doses (42 days if the first vaccination was with Vaxzevria)

**Table S4: Distribution of baseline characteristics** Absolute and relative frequencies.

| | 65-to-110-year-olds | 18-to-64-year-olds* |
|---|---|---|
| **Age in years** | | |
| 18–34 | 0 (0%) | 65006 (15%) |
| 35–49 | 0 (0%) | 121544 (27%) |
| 50–64 | 0 (0%) | 257777 (58%) |
| 65–79 | 890791 (74%) | 0 (0%) |
| 80–110 | 306710 (26%) | 0 (0%) |
| **Region of residence** | | |
| Helsinki-Uusimaa | 287190 (24%) | 130878 (29%) |
| Åland | 6691 (1%) | 1851 (0%) |
| Northern and Eastern Finland | 305550 (26%) | 108736 (24%) |
| Southern Finland | 282601 (24%) | 94503 (21%) |
| Western Finland | 315469 (26%) | 108359 (24%) |
| **Sex** | | |
| Female | 666298 (56%) | 232350 (52%) |
| Male | 531203 (44%) | 211977 (48%) |
| **Hospitalized in the year before the study** | | |
| No | 1042811 (87%) | 402995 (91%) |
| Yes | 154690 (13%) | 41332 (9%) |
| **Presence of comorbidities or medical therapies** | | |
| No predisposing medical conditions | 614246 (51%) | 0 (0%) |
| Only moderately predisposing medical conditions | 329422 (28%) | 316552 (71%) |
| At least one highly predisposing medical condition | 253833 (21%) | 127775 (29%) |
| **Long-term care** | | |
| No | 1161912 (97%) | 443910 (100%) |
| Yes | 35589 (3%) | 417 (0%) |
| **Number of vaccinations prior to the study** | | |
| 2 | 65707 (5%) | 88874 (20%) |
| 3 | 478171 (40%) | 296963 (67%) |
| 4–5 | 653623 (55%) | 58490 (13%) |
| **Last infection prior to the study** | | |
| No prior infection | 1115459 (93%) | 325893 (73%) |
| Pre-Omicron infection | 13314 (1%) | 14363 (3%) |
| Omicron infection | 68728 (6%) | 104071 (23%) |

* with underlying medical conditions

**Table S5: Hazard ratios, 65-to-110-year-olds** Covariate-adjusted hazard ratios comparing the hazards of severe COVID-19 outcomes and a negative control outcome in 65-to-110-year-olds who received a bivalent COVID-19 vaccine with the corresponding hazards in those who did not receive a bivalent COVID-19 vaccine.

| | | | Hazard ratio | | |
|---|---|---|---|---|---|
| Days since bivalent vaccination | Cases | Person-years | MLE | LCI | UCI |
| September – December 2022 | | | | | |
| **Hospitalisation due to COVID-19** | | | | | |
| No bivalent vaccination | 1471 | 227920 | | | |
| 0–13 | 71 | 22198 | 0.46 | 0.35 | 0.61 |
| 14–30 | 69 | 24199 | 0.41 | 0.31 | 0.55 |
| 31–60 | 59 | 23754 | 0.37 | 0.27 | 0.50 |
| 61–90 | 13 | 3159 | 0.66 | 0.38 | 1.17 |
| 91–120 | 0 | 94 | NE | NE | NE |
| **Death due to COVID-19** | | | | | |
| No bivalent vaccination | 870 | 228100 | | | |
| 0–13 | 15 | 22200 | 0.19 | 0.11 | 0.33 |
| 14–30 | 29 | 24204 | 0.34 | 0.23 | 0.51 |
| 31–60 | 44 | 23761 | 0.47 | 0.33 | 0.66 |
| 61–90 | 7–10 | 3160 | 0.50 | 0.25 | 0.99 |
| 91–120 | 1–4 | 95 | 3.34 | 0.82 | 13.62 |
| **Death in which COVID-19 was a contributing factor** | | | | | |
| No bivalent vaccination | 705 | 228100 | | | |
| 0–13 | 4–7 | 22200 | 0.11 | 0.05 | 0.23 |
| 14–30 | 31 | 24204 | 0.40 | 0.27 | 0.60 |
| 31–60 | 40 | 23761 | 0.45 | 0.31 | 0.64 |
| 61–90 | 7 | 3160 | 0.42 | 0.19 | 0.90 |
| 91–120 | 1–4 | 95 | 2.09 | 0.29 | 15.09 |
| **Emergency room visit due to injury** | | | | | |
| No bivalent vaccination | 7792 | 226899 | | | |
| 0–13 | 622 | 22077 | 0.86 | 0.78 | 0.95 |
| 14–30 | 689 | 24052 | 0.86 | 0.78 | 0.95 |
| 31–60 | 710 | 23597 | 0.92 | 0.84 | 1.02 |
| 61–90 | 88 | 3136 | 0.92 | 0.74 | 1.15 |
| 91–120 | 0 | 94 | NE | NE | NE |
| January – March 2023 | | | | | |
| **Hospitalisation due to COVID-19** | | | | | |
| No bivalent vaccination | 147 | 108817 | | | |
| 0–13 | 1–4 | 2506 | 1.23 | 0.45 | 3.38 |
| 14–30 | 11 | 5663 | 1.24 | 0.65 | 2.38 |
| 31–60 | 32 | 28213 | 0.71 | 0.46 | 1.12 |
| 61–90 | 51 | 46267 | 0.95 | 0.65 | 1.40 |
| 91–120 | 38 | 43788 | 0.78 | 0.51 | 1.19 |
| 121–150 | 41 | 23061 | 1.23 | 0.79 | 1.91 |
| 151–180 | 6–9 | 2976 | 1.21 | 0.51 | 2.86 |
| 181–211 | 0 | 86 | NE | NE | NE |
| **Death due to COVID-19** | | | | | |
| No bivalent vaccination | 99 | 108872 | | | |
| 0–13 | 0 | 2506 | NE | NE | NE |
| 14–30 | 2–5 | 5664 | 0.81 | 0.32 | 2.05 |
| 31–60 | 29 | 28222 | 0.86 | 0.52 | 1.41 |

(Table S5 continued)

| Days since bivalent vaccination | Cases | Person-years | MLE | LCI | UCI |
|---|---|---|---|---|---|
| 61–90 | 35 | 46280 | 0.85 | 0.54 | 1.35 |
| 91–120 | 18 | 43796 | 0.75 | 0.42 | 1.33 |
| 121–150 | 11 | 23065 | 0.90 | 0.44 | 1.87 |
| 151–180 | 1–4 | 2977 | 0.50 | 0.07 | 3.80 |
| 181–211 | 0 | 86 | NE | NE | NE |
| **Death in which COVID-19 was a contributing factor** | | | | | |
| No bivalent vaccination | 100 | 108872 | | | |
| 0–13 | 1–4 | 2506 | 0.30 | 0.04 | 2.19 |
| 14–30 | 1–4 | 5664 | 0.41 | 0.15 | 1.14 |
| 31–60 | 22 | 28222 | 0.40 | 0.24 | 0.67 |
| 61–90 | 28 | 46280 | 0.41 | 0.25 | 0.65 |
| 91–120 | 25 | 43796 | 0.82 | 0.49 | 1.37 |
| 121–150 | 13 | 23065 | 1.30 | 0.63 | 2.66 |
| 151–180 | 0 | 2977 | NE | NE | NE |
| 181–211 | 0 | 86 | NE | NE | NE |
| **Emergency room visit due to injury** | | | | | |
| No bivalent vaccination | 2746 | 107540 | | | |
| 0–13 | 68–71 | 2478 | 1.12 | 0.88 | 1.42 |
| 14–30 | 153 | 5599 | 1.07 | 0.90 | 1.26 |
| 31–60 | 724 | 27926 | 0.97 | 0.88 | 1.07 |
| 61–90 | 1194 | 45779 | 0.97 | 0.89 | 1.05 |
| 91–120 | 1103 | 43272 | 0.95 | 0.88 | 1.04 |
| 121–150 | 605 | 22775 | 0.93 | 0.83 | 1.03 |
| 151–180 | 79 | 2938 | 0.88 | 0.70 | 1.11 |
| 181–211 | 1–4 | 85 | 1.12 | 0.36 | 3.50 |

MLE, maximum likelihood estimate;

LCI/UCI, lower/upper limit of the 95% Wald confidence interval;

NE, not estimated

**Table S6: Hazard ratios, 65-to-79-year-olds** Covariate-adjusted hazard ratios comparing the hazards of severe COVID-19 outcomes in 65-to-79-year-olds who received a bivalent COVID-19 vaccine with the corresponding hazards in those who did not receive a bivalent COVID-19 vaccine.

| | | | Hazard ratio | | |
|---|---|---|---|---|---|
| Days since bivalent vaccination | Cases | Person-years | MLE | LCI | UCI |
| September – December 2022 | | | | | |
| **Hospitalisation due to COVID-19** | | | | | |
| No bivalent vaccination | 626 | 156640 | | | |
| 0–13 | 25 | 15707 | 0.41 | 0.26 | 0.63 |
| 14–60 | 72 | 33421 | 0.55 | 0.40 | 0.75 |
| 61–120 | 7 | 2283 | 0.83 | 0.38 | 1.79 |
| **Death due to COVID-19** | | | | | |
| No bivalent vaccination | 190 | 156721 | | | |
| 0–13 | 7 | 15707 | 0.43 | 0.19 | 0.97 |
| 14–60 | 15 | 33426 | 0.44 | 0.24 | 0.81 |
| 61–120 | ND | 2284 | 0.60 | 0.14 | 2.56 |
| **Death in which COVID-19 was a contributing factor** | | | | | |
| No bivalent vaccination | 172 | 156721 | | | |
| 0–13 | ND | 15707 | 0.07 | 0.01 | 0.54 |
| 14–60 | 14 | 33426 | 0.44 | 0.23 | 0.86 |
| 61–120 | ND | 2284 | 0.65 | 0.15 | 2.74 |
| January – March 2023 | | | | | |
| **Hospitalisation due to COVID-19** | | | | | |
| No bivalent vaccination | 82 | 85025 | | | |
| 0–13 | ND | 1989 | 0.54 | 0.07 | 3.93 |
| 14–60 | 24 | 25251 | 0.90 | 0.52 | 1.55 |
| 61–120 | 49 | 64282 | 0.95 | 0.60 | 1.49 |
| 121–180 | ND | 18181 | 1.80 | 1.03 | 3.14 |
| 181–211 | 0 | 67 | NE | NE | NE |
| **Death due to COVID-19** | | | | | |
| No bivalent vaccination | 33 | 85053 | | | |
| 0–13 | 0 | 1989 | NE | NE | NE |
| 14–60 | ND | 25256 | 0.42 | 0.17 | 1.04 |
| 61–120 | 9 | 64295 | 0.33 | 0.14 | 0.78 |
| 121–180 | ND | 18184 | 0.45 | 0.11 | 1.77 |
| 181–211 | 0 | 67 | NE | NE | NE |
| **Death in which COVID-19 was a contributing factor** | | | | | |
| No bivalent vaccination | 30 | 85053 | | | |
| 0–13 | ND | 1989 | 0.93 | 0.12 | 7.06 |
| 14–60 | ND | 25256 | 0.63 | 0.27 | 1.46 |
| 61–120 | 10 | 64295 | 0.37 | 0.16 | 0.86 |
| 121–180 | 7 | 18184 | 2.08 | 0.71 | 6.08 |
| 181–211 | 0 | 67 | NE | NE | NE |

MLE, maximum likelihood estimate;

LCI/UCI, lower/upper limit of the 95% Wald confidence interval;

ND, not disclosed;

NE, not estimated

**Table S7: Hazard ratios, 80-to-110-year-olds** Covariate-adjusted hazard ratios comparing the hazards of severe COVID-19 outcomes in 80-to-110-year-olds who received a bivalent COVID-19 vaccine with the corresponding hazards in those who did not receive a bivalent COVID-19 vaccine.

| | | | Hazard ratio | | |
|---|---|---|---|---|---|
| Days since bivalent vaccination | Cases | Person-years | MLE | LCI | UCI |
| September – December 2022 | | | | | |
| **Hospitalisation due to COVID-19** | | | | | |
| No bivalent vaccination | 845 | 71280 | | | |
| 0–13 | 46 | 6491 | 0.48 | 0.34 | 0.69 |
| 14–60 | 56 | 14533 | 0.28 | 0.20 | 0.39 |
| 61–120 | 6 | 970 | 0.53 | 0.23 | 1.21 |
| **Death due to COVID-19** | | | | | |
| No bivalent vaccination | 680 | 71380 | | | |
| 0–13 | 8 | 6493 | 0.13 | 0.06 | 0.27 |
| 14–60 | 58 | 14539 | 0.39 | 0.28 | 0.55 |
| 61–120 | ND | 971 | 0.58 | 0.29 | 1.16 |
| **Death in which COVID-19 was a contributing factor** | | | | | |
| No bivalent vaccination | 533 | 71380 | | | |
| 0–13 | ND | 6493 | 0.12 | 0.05 | 0.27 |
| 14–60 | 57 | 14539 | 0.41 | 0.30 | 0.58 |
| 61–120 | ND | 971 | 0.42 | 0.18 | 0.97 |
| January – March 2023 | | | | | |
| **Hospitalisation due to COVID-19** | | | | | |
| No bivalent vaccination | 65 | 23792 | | | |
| 0–13 | ND | 518 | 2.09 | 0.63 | 6.90 |
| 14–60 | 19 | 8625 | 0.67 | 0.36 | 1.25 |
| 61–120 | 40 | 25773 | 0.80 | 0.48 | 1.33 |
| 121–180 | ND | 7856 | 0.81 | 0.42 | 1.55 |
| 181–211 | 0 | 20 | NE | NE | NE |
| **Death due to COVID-19** | | | | | |
| No bivalent vaccination | 66 | 23819 | | | |
| 0–13 | 0 | 518 | NE | NE | NE |
| 14–60 | ND | 8630 | 1.10 | 0.63 | 1.91 |
| 61–120 | 44 | 25782 | 1.10 | 0.69 | 1.78 |
| 121–180 | ND | 7858 | 1.12 | 0.49 | 2.56 |
| 181–211 | 0 | 20 | NE | NE | NE |
| **Death in which COVID-19 was a contributing factor** | | | | | |
| No bivalent vaccination | 70 | 23819 | | | |
| 0–13 | 0 | 518 | NE | NE | NE |
| 14–60 | ND | 8630 | 0.34 | 0.18 | 0.62 |
| 61–120 | 43 | 25782 | 0.57 | 0.36 | 0.91 |
| 121–180 | 6 | 7858 | 0.65 | 0.24 | 1.74 |
| 181–211 | 0 | 20 | NE | NE | NE |

MLE, maximum likelihood estimate;

LCI/UCI, lower/upper limit of the 95% Wald confidence interval;

ND, not disclosed;

NE, not estimated

**Table S8: Hazard ratios, 65-to-110-year-olds, stratified by vaccine composition** Covariate-adjusted hazard ratios comparing the hazards of severe COVID-19 outcomes in 65-to-110-year-olds who received a bivalent COVID-19 vaccine with the corresponding hazards in those who did not receive a bivalent COVID-19 vaccine distinguishing between BA.1 and BA.4-5 bivalent vaccines.

| | | | Hazard ratio | | |
|---|---|---|---|---|---|
| Days since bivalent vaccination | Cases | Person-years | MLE | LCI | UCI |
| September – December 2022 | | | | | |
| **Hospitalisation due to COVID-19** | | | | | |
| No bivalent vaccination | 1471 | 227920 | | | |
| *BA.1 bivalent vaccine* | | | | | |
| 0–13 | 20 | 6970 | 0.38 | 0.24 | 0.61 |
| 14–60 | 39 | 15422 | 0.34 | 0.24 | 0.48 |
| 61–120 | 9–12 | 1746 | 0.88 | 0.46 | 1.66 |
| *BA.4-5 bivalent vaccine* | | | | | |
| 0–13 | 51 | 15228 | 0.50 | 0.37 | 0.68 |
| 14–60 | 89 | 32531 | 0.42 | 0.33 | 0.55 |
| 61–120 | 1–4 | 1507 | 0.35 | 0.11 | 1.10 |
| **Death due to COVID-19** | | | | | |
| No bivalent vaccination | 870 | 228100 | | | |
| *BA.1 bivalent vaccine* | | | | | |
| 0–13 | 1–4 | 6971 | 0.17 | 0.06 | 0.46 |
| 14–60 | 16 | 15426 | 0.29 | 0.17 | 0.48 |
| 61–120 | ND | 1747 | 0.80 | 0.35 | 1.82 |
| *BA.4-5 bivalent vaccine* | | | | | |
| 0–13 | 11–14 | 15229 | 0.21 | 0.11 | 0.38 |
| 14–60 | 57 | 32540 | 0.46 | 0.34 | 0.63 |
| 61–120 | ND | 1508 | 0.44 | 0.18 | 1.09 |
| **Death in which COVID-19 was a contributing factor** | | | | | |
| No bivalent vaccination | 705 | 228100 | | | |
| *BA.1 bivalent vaccine* | | | | | |
| 0–13 | ND | 6971 | 0.21 | 0.08 | 0.57 |
| 14–60 | 18 | 15426 | 0.36 | 0.22 | 0.60 |
| 61–120 | ND | 1747 | 0.58 | 0.21 | 1.58 |
| *BA.4-5 bivalent vaccine* | | | | | |
| 0–13 | ND | 15229 | 0.07 | 0.02 | 0.21 |
| 14–60 | 53 | 32540 | 0.45 | 0.33 | 0.63 |
| 61–120 | ND | 1508 | 0.38 | 0.14 | 1.04 |
| January – March 2023 | | | | | |
| **Hospitalisation due to COVID-19** | | | | | |
| No bivalent vaccination | 147 | 108817 | | | |
| *BA.1 bivalent vaccine* | | | | | |
| 0–13 | ND | 479 | 1.62 | 0.22 | 11.70 |
| 14–60 | 5 | 9410 | 0.34 | 0.13 | 0.85 |
| 61–120 | 16 | 27420 | 0.55 | 0.31 | 0.96 |
| 121–180 | ND | 9039 | 0.95 | 0.49 | 1.85 |
| *BA.4-5 bivalent vaccine* | | | | | |
| 0–13 | ND | 2027 | 1.13 | 0.35 | 3.59 |
| 14–60 | 38 | 24466 | 0.96 | 0.63 | 1.47 |
| 61–120 | 73 | 62635 | 1.02 | 0.71 | 1.45 |

(Table S8 continued)

| Days since bivalent vaccination | Cases | Person-years | MLE | LCI | UCI |
|---|---|---|---|---|---|
| 121–180 | ND | 16998 | 1.45 | 0.92 | 2.30 |
| *BA.1 and BA.4-5 bivalent vaccines* | | | | | |
| 181–211 | 0 | 86 | NE | NE | NE |
| **Death due to COVID-19** | | | | | |
| No bivalent vaccination | 99 | 108872 | | | |
| *BA.1 bivalent vaccine* | | | | | |
| 0–13 | 0 | 479 | NE | NE | NE |
| 14–60 | ND | 9412 | 0.84 | 0.41 | 1.70 |
| 61–120 | 20 | 27425 | 1.07 | 0.62 | 1.85 |
| 121–180 | ND | 9041 | 0.94 | 0.32 | 2.71 |
| *BA.4-5 bivalent vaccine* | | | | | |
| 0–13 | 0 | 2027 | NE | NE | NE |
| 14–60 | ND | 24473 | 0.85 | 0.51 | 1.43 |
| 61–120 | 33 | 62651 | 0.70 | 0.44 | 1.12 |
| 121–180 | ND | 17001 | 0.82 | 0.36 | 1.87 |
| *BA.1 and BA.4-5 bivalent vaccines* | | | | | |
| 181–211 | 0 | 86 | NE | NE | NE |
| **Death in which COVID-19 was a contributing factor** | | | | | |
| No bivalent vaccination | 100 | 108872 | | | |
| *BA.1 bivalent vaccine* | | | | | |
| 0–13 | ND | 479 | 1.55 | 0.21 | 11.24 |
| 14–60 | ND | 9412 | 0.70 | 0.36 | 1.34 |
| 61–120 | 18 | 27425 | 0.71 | 0.41 | 1.23 |
| 121–180 | 1–4 | 9041 | 1.09 | 0.37 | 3.17 |
| *BA.4-5 bivalent vaccine* | | | | | |
| 0–13 | 0 | 2027 | NE | NE | NE |
| 14–60 | ND | 24473 | 0.31 | 0.17 | 0.57 |
| 61–120 | 35 | 62651 | 0.46 | 0.29 | 0.72 |
| 121–180 | 9–12 | 17001 | 0.98 | 0.43 | 2.21 |
| *BA.1 and BA.4-5 bivalent vaccines* | | | | | |
| 181–211 | 0 | 86 | NE | NE | NE |

MLE, maximum likelihood estimate;

LCI/UCI, lower/upper limit of the 95% Wald confidence interval;

ND, not disclosed;

NE, not estimated

**Table S9: Hazard ratios, 18-to-64-year-olds with underlying medical conditions** Covariate-adjusted hazard ratios comparing the hazards of severe COVID-19 outcomes and a negative control outcome in 18-to-64-year-olds with underlying medical conditions who received a bivalent COVID-19 vaccine with the corresponding hazards in those who did not receive a bivalent COVID-19 vaccine.

| | | | Hazard ratio | | |
|---|---|---|---|---|---|
| Days since bivalent vaccination | Cases | Person-years | MLE | LCI | UCI |
| September – December 2022 | | | | | |
| **Hospitalisation due to COVID-19** | | | | | |
| No bivalent vaccination | 204 | 119381 | | | |
| 0–13 | 1–4 | 2231 | 0.16 | 0.02 | 1.16 |
| 14–30 | 6 | 2367 | 1.01 | 0.43 | 2.40 |
| 31–60 | 9 | 2408 | 1.81 | 0.87 | 3.76 |
| 61–90 | 1–4 | 517 | 3.82 | 1.17 | 12.43 |
| 91–120 | 0 | 21 | NE | NE | NE |
| **Death due to COVID-19** | | | | | |
| No bivalent vaccination | 14 | 119406 | | | |
| 0–13 | 0 | 2231 | NE | NE | NE |
| 14–30 | 0 | 2367 | NE | NE | NE |
| 31–60 | 0 | 2408 | NE | NE | NE |
| 61–90 | 0 | 517 | NE | NE | NE |
| 91–120 | 0 | 21 | NE | NE | NE |
| **Death in which COVID-19 was a contributing factor** | | | | | |
| No bivalent vaccination | 14 | 119406 | | | |
| 0–13 | 0 | 2231 | NE | NE | NE |
| 14–30 | 1–4 | 2367 | 2.23 | 0.24 | 20.88 |
| 31–60 | 1–4 | 2408 | 2.31 | 0.24 | 22.22 |
| 61–90 | 0 | 517 | NE | NE | NE |
| 91–120 | 0 | 21 | NE | NE | NE |
| **Emergency room visit due to injury** | | | | | |
| No bivalent vaccination | 2085 | 119057 | | | |
| 0–13 | 56 | 2222 | 1.51 | 1.14 | 2.00 |
| 14–30 | 54 | 2356 | 1.40 | 1.05 | 1.87 |
| 31–60 | 37 | 2396 | 1.00 | 0.71 | 1.41 |
| 61–90 | 15 | 514 | 2.02 | 1.21 | 3.40 |
| 91–120 | 0 | 20 | NE | NE | NE |
| January – March 2023 | | | | | |
| **Hospitalisation due to COVID-19** | | | | | |
| No bivalent vaccination | 35 | 81328 | | | |
| 0–13 | 0 | 499 | NE | NE | NE |
| 14–30 | 1–4 | 922 | 1.32 | 0.17 | 10.05 |
| 31–60 | 2 | 3217 | 0.74 | 0.16 | 3.30 |
| 61–90 | 7 | 4644 | 2.60 | 1.04 | 6.48 |
| 91–120 | 7 | 4319 | 3.40 | 1.34 | 8.66 |
| 121–150 | 2 | 2321 | 2.05 | 0.44 | 9.51 |
| 151–180 | 1–4 | 486 | 5.33 | 0.66 | 42.76 |
| 181–211 | 0 | 19 | NE | NE | NE |
| **Death due to COVID-19** | | | | | |
| No bivalent vaccination | 1–4 | 81339 | | | |
| 0–13 | 0 | 499 | NE | NE | NE |
| 14–30 | 0 | 922 | NE | NE | NE |

(Table S9 continued)

| Days since bivalent vaccination | Cases | Person-years | MLE | LCI | UCI |
|---|---|---|---|---|---|
| 31–60 | 1–4 | 3218 | NE | NE | NE |
| 61–90 | 0 | 4645 | NE | NE | NE |
| 91–120 | 0 | 4320 | NE | NE | NE |
| 121–150 | 0 | 2321 | NE | NE | NE |
| 151–180 | 0 | 486 | NE | NE | NE |
| 181–211 | 0 | 19 | NE | NE | NE |
| **Death in which COVID-19 was a contributing factor** | | | | | |
| No bivalent vaccination | 1–4 | 81339 | | | |
| 0–13 | 0 | 499 | NE | NE | NE |
| 14–30 | 0 | 922 | NE | NE | NE |
| 31–60 | 1–4 | 3218 | NE | NE | NE |
| 61–90 | 0 | 4645 | NE | NE | NE |
| 91–120 | 0 | 4320 | NE | NE | NE |
| 121–150 | 0 | 2321 | NE | NE | NE |
| 151–180 | 0 | 486 | NE | NE | NE |
| 181–211 | 0 | 19 | NE | NE | NE |
| **Emergency room visit due to injury** | | | | | |
| No bivalent vaccination | 1244 | 80729 | | | |
| 0–13 | 6 | 495 | 0.80 | 0.36 | 1.78 |
| 14–30 | 16 | 915 | 1.14 | 0.69 | 1.88 |
| 31–60 | 61 | 3194 | 1.21 | 0.92 | 1.59 |
| 61–90 | 76 | 4608 | 1.11 | 0.87 | 1.42 |
| 91–120 | 75 | 4281 | 1.20 | 0.93 | 1.53 |
| 121–150 | 34 | 2299 | 0.96 | 0.67 | 1.37 |
| 151–180 | 6 | 480 | 0.76 | 0.34 | 1.71 |
| 181–211 | 0 | 19 | NE | NE | NE |

MLE, maximum likelihood estimate;

LCI/UCI, lower/upper limit of the 95% Wald confidence interval;

NE, not estimated

**Figure S1: Distribution of sequenced Omicron lineages in Finland** The graph shows the percentages of the most frequent SARS-CoV-2 sublineages detected by sentinel surveillance and recorded in the National Infectious Diseases Register by week of sample collection since August 2022. Green bars represent BA.2 sublineages (BA.2 lineages, BA.2.75.X and CH.1.1.X), orange bars represent BA.4 sublineages and pink bars represent BA.5 sublineages (BA.5 lineages, BF.7.X and BQ.1.X). The grey bars represent recombinant lineages (XBB, XBB.1.5, XBB.1.9.1.X, XBB.1.16.X and XBF) as well as the group 'Other variants', which comprises all sublineages not assigned to any of the previous groups. The .X extension covers the whole sublineage family, e.g. BQ.1.X covers all descendants of BQ.1, such as BQ.1.1 and BQ.1.2.

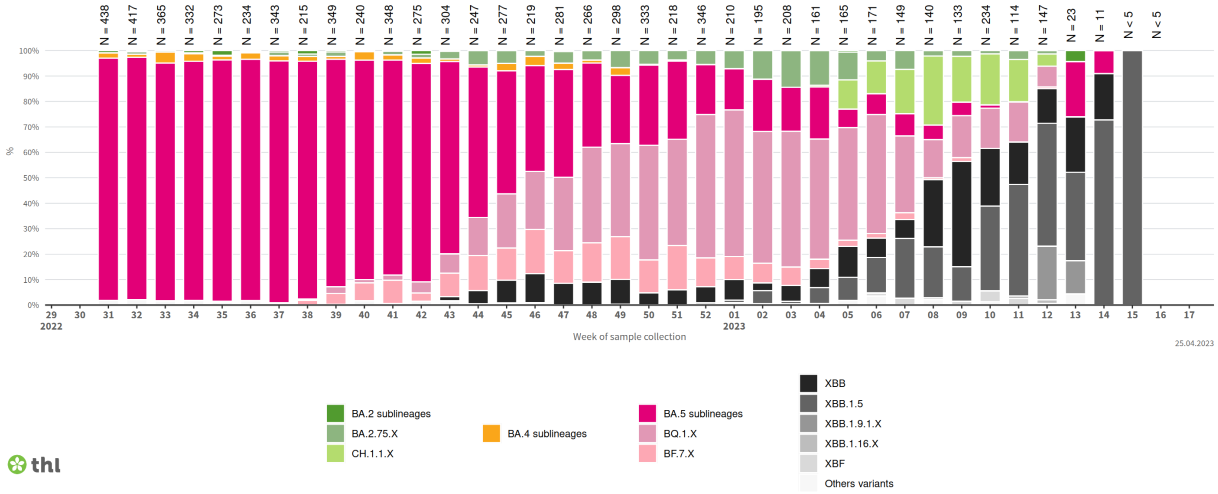

Adapted from https://thl.fi/en/web/infectious-diseases-and-vaccinations/what-s-new/coronavirus-covid-19-latest-updates/coronavirus-variants/genomic-surveillance-of-sars-cov-2
