## Supplementary material for "Bivalent booster effectiveness against severe COVID-19 outcomes in Finland, September 2022 – March 2023": Ethical statement

To whom it may concern:

As director of the department for Health security of the Finnish Institute for Health and Welfare, I certify that:

- I am the competent auhority for assessing whether research requires institutionnal ethical review or if the Finnish communicable diseases law (Tartuntatautilaki 1227/2016) and the law on the duties of the Finnish institute for Health (Laki Terveyden ja hyvinvoinnin laitoksesta 668/2008) and Welfare allows the implementation of the research without seeking further ethical review.

- The research presented by Poukka et al in "Bivalent booster effectiveness against severe COVID-19 outcomes in Finland, September 2022 – March 2023" did not require further ethical review before implementation as its aim was to monitor vaccine effectiveness of infectious disease (Tartuntatautilaki 1227/2016).

Helsinki, May 5th 2023
Chief Doctor Otto Helve
